# Sub-5-minute Detection of SARS-CoV-2 RNA using a Reverse Transcriptase-Free Exponential Amplification Reaction, RTF-EXPAR

**DOI:** 10.1101/2020.12.31.20248236

**Authors:** Jake G. Carter, Lorea Orueta Iturbe, Jean-Louis H. A. Duprey, Ian R. Carter, Craig D. Southern, Marium Rana, Andrew Bosworth, Andrew D. Beggs, Matthew R. Hicks, James H. R. Tucker, Timothy R. Dafforn

## Abstract

We report a rapid isothermal method for detecting SARS-CoV-2, the virus responsible for COVID-19. The procedure uses a novel reverse transcriptase-free (RTF) approach for converting RNA into DNA, which triggers a rapid amplification using the Exponential Amplification Reaction (EXPAR). Deploying the RNA-to-DNA conversion and amplification stages of the RTF-EXPAR assay in a single step results in the detection of a sample of patient SARS-CoV-2 RNA in under 5 minutes.

---

In order to reduce the rate of spread of COVID-19, an accurate and efficient virus testing strategy is imperative. A key part of this strategy is continuous assay development, with the aim of reducing detection times and increasing sample throughput. The research community and diagnostics industry has responded rapidly to this unprecedented crisis in developing a range of detection platforms.^1-4^ The most sensitive assays detect viral RNA, with the current gold standard being reverse transcriptase polymerase chain reaction (RT-PCR), a two-step assay that takes more than 60 minutes per sample. First, reverse transcriptase converts viral RNA to complementary DNA (cDNA), a process that can take up to 30 minutes.^5^ Then a quantitative PCR (qPCR) amplifies the cDNA, which is detected using a fluorescent dye, a process that takes up to an hour.^6^ To reduce assay times, a plethora of new approaches to SARS-CoV-2 detection have appeared in the literature over the past year. As far as Nucleic Acid Amplification Tests (NAATs) are concerned, which are more sensitive than current 30- minute lateral flow antigen (immunoassay) tests,^7^ focus has turned towards isothermal DNA amplification approaches, which increase amplification speeds and hence reduce assay times. The most common isothermal amplification system is Loop mediated isothermal AMPlification (LAMP).^8^ LAMP assays have been developed for SARS-CoV-2 but take, on average, 20 minutes for a result, with further decreases in LAMP assay time proving challenging.^4,9,10^ Herein, we demonstrate an alternative isothermal approach based on the Exponential Amplification Reaction (EXPAR),^11^ a simpler and faster amplification method than LAMP. By combining EXPAR with a novel reverse transcriptase-free (RTF) step, this new assay, RTF-EXPAR, can accurately identify viral RNA derived from COVID-19 patient samples in less than 5 minutes.

The key to the speed of EXPAR is twofold; firstly, the amplification occurs at a single temperature, thus avoiding lengthy heating and cooling steps, and secondly the amplicon is relatively small (typically 15-20 bases long), compared to both PCR and LAMP. These two factors result in EXPAR, once triggered, producing up to 10^8^ strands of DNA product in a matter of minutes.^11,12^ A single-stranded DNA fragment (the trigger) starts the EXPAR reaction by binding a DNA template. Large quantities of short double stranded DNA sequences are then generated in an isothermal cycle involving a DNA polymerase to extend the sequence and a nicking endonuclease to cut it, while leaving the template intact (Scheme 1a). As with the RT-PCR COVID-19 assay, duplex formation is monitored spectroscopically using a fluorescent intercalating dye, e.g. SYBR Green.

A crucial element to developing a successful EXPAR assay is the identification of optimal nucleotide sequences in the target genome. Qian *et al* previously found that the type of trigger sequence used in EXPAR plays a vital role in determining its efficiency.^13,14^ Using their approach, we designed a 17- mer DNA trigger for EXPAR (**Trigger X**, Scheme 1a and Table 1) containing a sequence complementary to one within the conserved gene *Orf1ab* in the SARS-CoV-2 genome (https://www.ncbi.nlm.nih.gov/nuccore/MN908947.3?report=fasta). We first analysed the speed and sensitivity of EXPAR using **Trigger X** in the presence of **Template X’-X’** (Supplementary Figure 1). Rapid rises in SYBR Green fluorescence were observed, with amplification times revealing an expected dependence on trigger concentration (e.g. time to 10-sigma: 3.17 ± 0.14 minutes at 10 nM, 8.67 ± 1.08 minutes at 10 pM). These results demonstrate that EXPAR is a faster amplification method than LAMP. Next, we analysed the specificity of the reaction by investigating three other triggers (**Triggers A, B** and **C**), each at a concentration of 10 nM, that were non-complementary to **Template X’-X’** (Supplementary Figure 2). Each of these three triggers produced no signal within 10 minutes under the same conditions, confirming the specificity of the EXPAR reaction, with only the trigger sequence fully complementary to the template (**Trigger X**) resulting in rapid amplification.

**Table 1.**
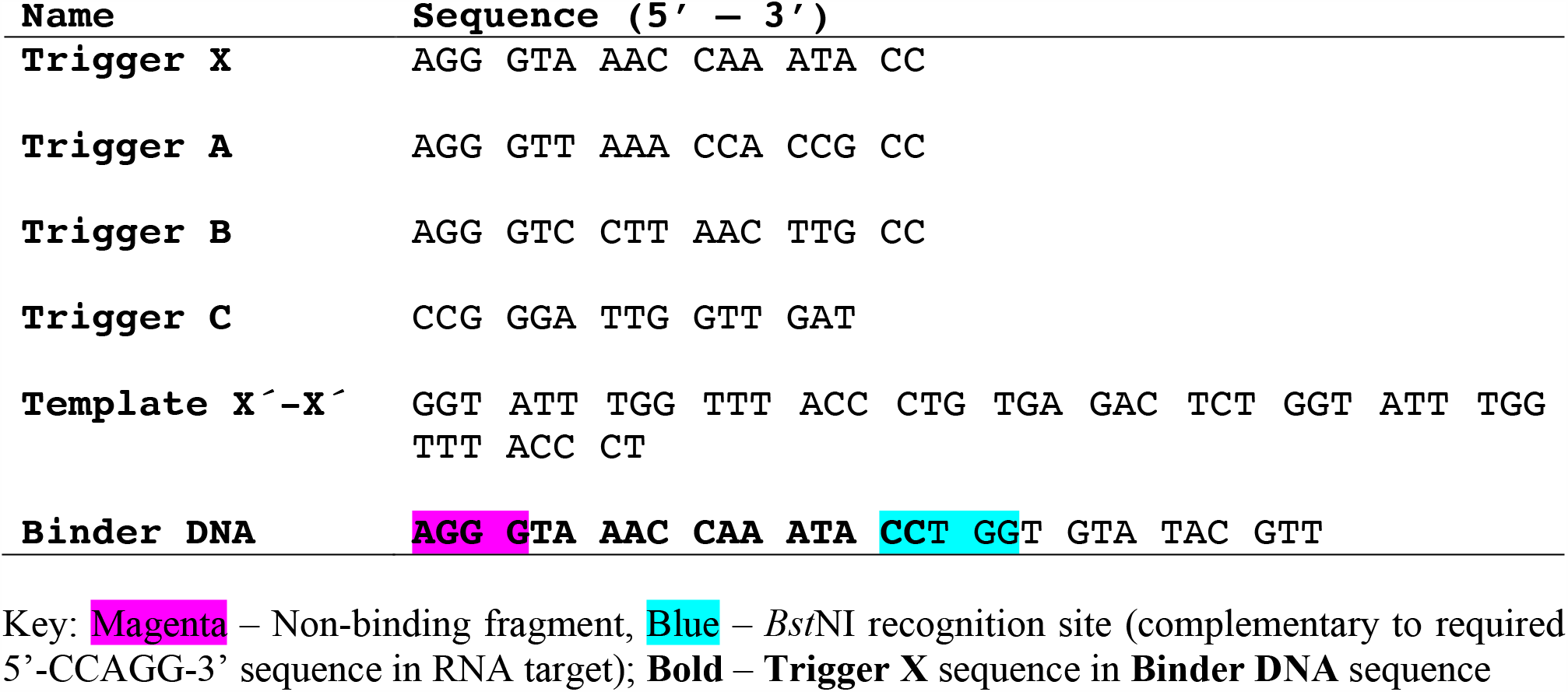
Oligonucleotides used in study.

In the standard RT-PCR COVID-19 assay, reverse transcriptase converts the RNA of SARS-CoV-2 into cDNA prior to amplification (*vide supra*). The speed of this initial polymerisation reaction is a significant limitation for this or potentially any other RNA detection method that proceeds via cDNA amplification, including LAMP or EXPAR. We hypothesised that a faster method could be achieved by generating a short DNA trigger sequence directly from the RNA genomic strand, without the need for a lengthy reverse transcriptase step. Murray *et al* had previously demonstrated that the restriction enzyme *Bst*NI could act as a nicking enzyme by selectively cleaving DNA within RNA:DNA heteroduplexes.^15^ We considered that this enzyme could be used to generate the desired DNA fragment for triggering the EXPAR reaction. To achieve this, we designed a 30-mer oligonucleotide (called **Binder DNA**, Table 1) possessing a 5-base recognition site for *Bst*NI, as well as two partially overlapping sequence stretches complementary to part of *Orf1ab* in the SARS-CoV-2 RNA genome and the EXPAR DNA template (**Template X’-X’**). Site-selective cleavage of **Binder DNA** using *Bst*NI would only occur in the presence of the RNA target from SARS-CoV-2, generating a shorter strand of DNA, **Trigger X** (Scheme 1b). This shorter strand would now release from the heteroduplex and bind preferably to the DNA template, as it can still form a fully-complementary 17-mer duplex with the latter. Binding to the template would trigger EXPAR, with the newly released RNA strand able to bind more **Binder DNA** to generate more **Trigger X**.

Applying this novel EXPAR approach in a two-stage process, we first undertook an enzymatic digestion at 50 °C for five minutes of **Binder DNA** (1 µM) in the presence of a sample of patient SARS-CoV-2 RNA (72.7 copies/µL)^16^ obtained from PHE, before adding this solution to the EXPAR reagent mix for the amplification step. This stage, performed in triplicate, gave an amplification time of 3.17 ± 0.24 minutes, whereas no amplification was observed for the negative sample within 10 minutes (Fig. 1 and Supplementary Figure 3). To increase the speed of the RTF-EXPAR assay further, we next investigated a “one-pot” approach by introducing *Bst*NI and **Binder DNA** to the EXPAR reagents at the same time, before incubating and amplifying simultaneously at 50 °C. These assay conditions gave an amplification time of only 4.00 ± 0.72 minutes for the positive sample, halving the total assay time compared to the “two-pot” method (Fig. 1 and Supplementary Figure 4). Once again, no signal change for the negative sample was observed within 10 minutes. As expected, this was also the case for control experiments on the positive RNA sample in the absence of either **Binder DNA** or **Template X’-X’** (see Supplementary Figures 5 and 6 respectively), and a sample of RNA isolated from the CFPAC-1 human ductal pancreatic adenocarcinoma cell line (see Supplementary Figure 7).

**Figure 1.**
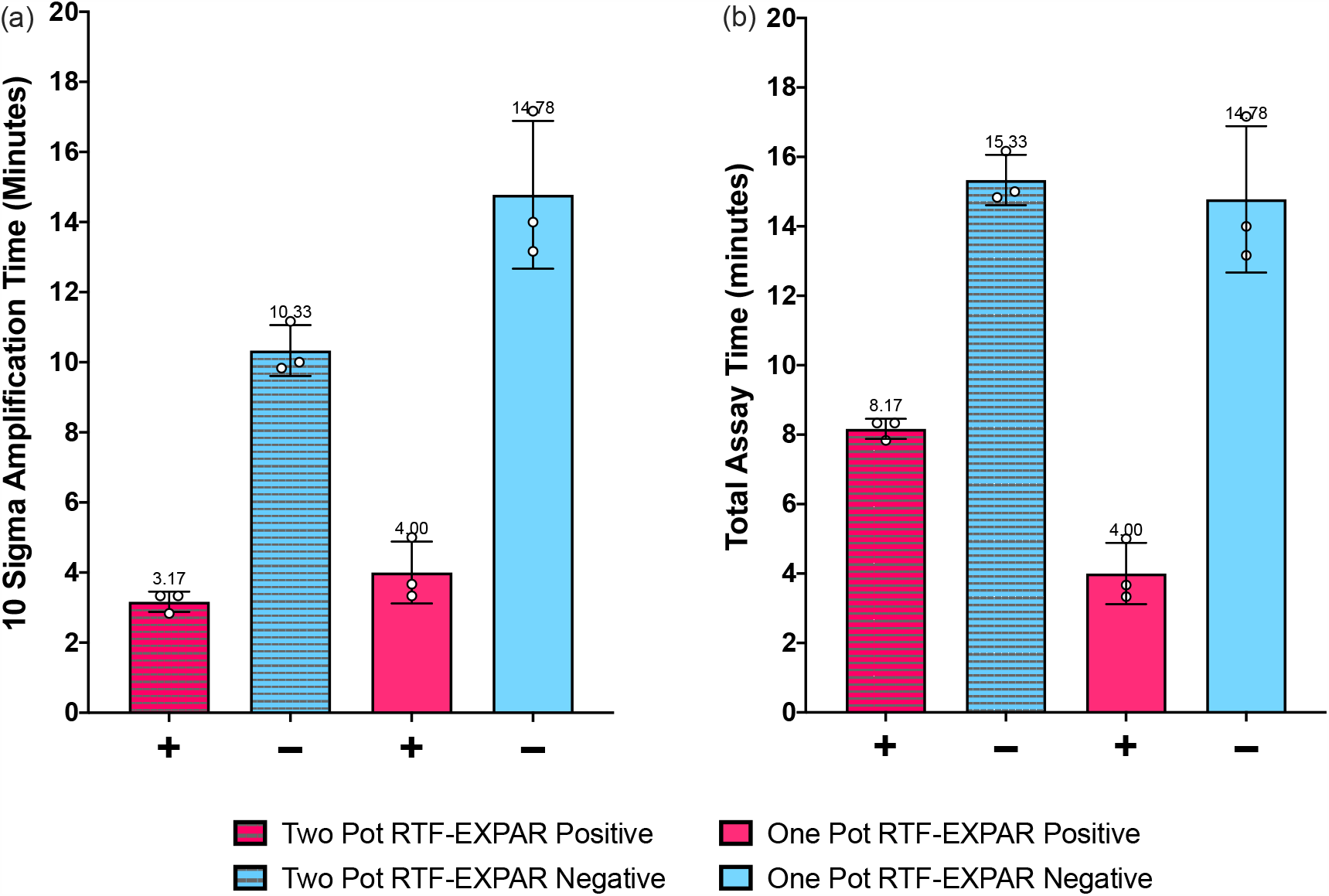
RTF-EXPAR assay data for SARS-CoV-2 RNA detection (72.7 copies/µL, n = 3), showing: (a) the mean time for the amplification reaction only and (b) the mean total assay time from RNA sample to signal. Each run time was calculated to be the point at which the fluorescence signal was greater than 10 standard deviations from the baseline signal (10-sigma time). Error bars in datasets are the standard deviations of the 10-sigma time. Signals observed for negative samples at >10 min are attributed to amplification arising from non-specific interactions.

In conclusion, through the use of a new reverse transcriptase-free isothermal amplification method, RTF-EXPAR, involving a DNA-selective restriction endonuclease, we have demonstrated the successful detection of SARS-CoV-2 RNA in a total assay time of less than 5 minutes. This time is not only much faster than RT-PCR (assay time of at least 60 minutes) but also outperforms LAMP and 30-minute lateral flow antigen tests in current deployment. RTF-EXPAR would be completely compatible (i.e. deployment ready) for use on equipment currently used for RT-PCR COVID-19 assays. Furthermore, the simplicity and speed of the assay enables this method to be modified to detect a range of infectious diseases caused by RNA-based pathogens (e.g. Ebola, RSV).

**Scheme 1.**
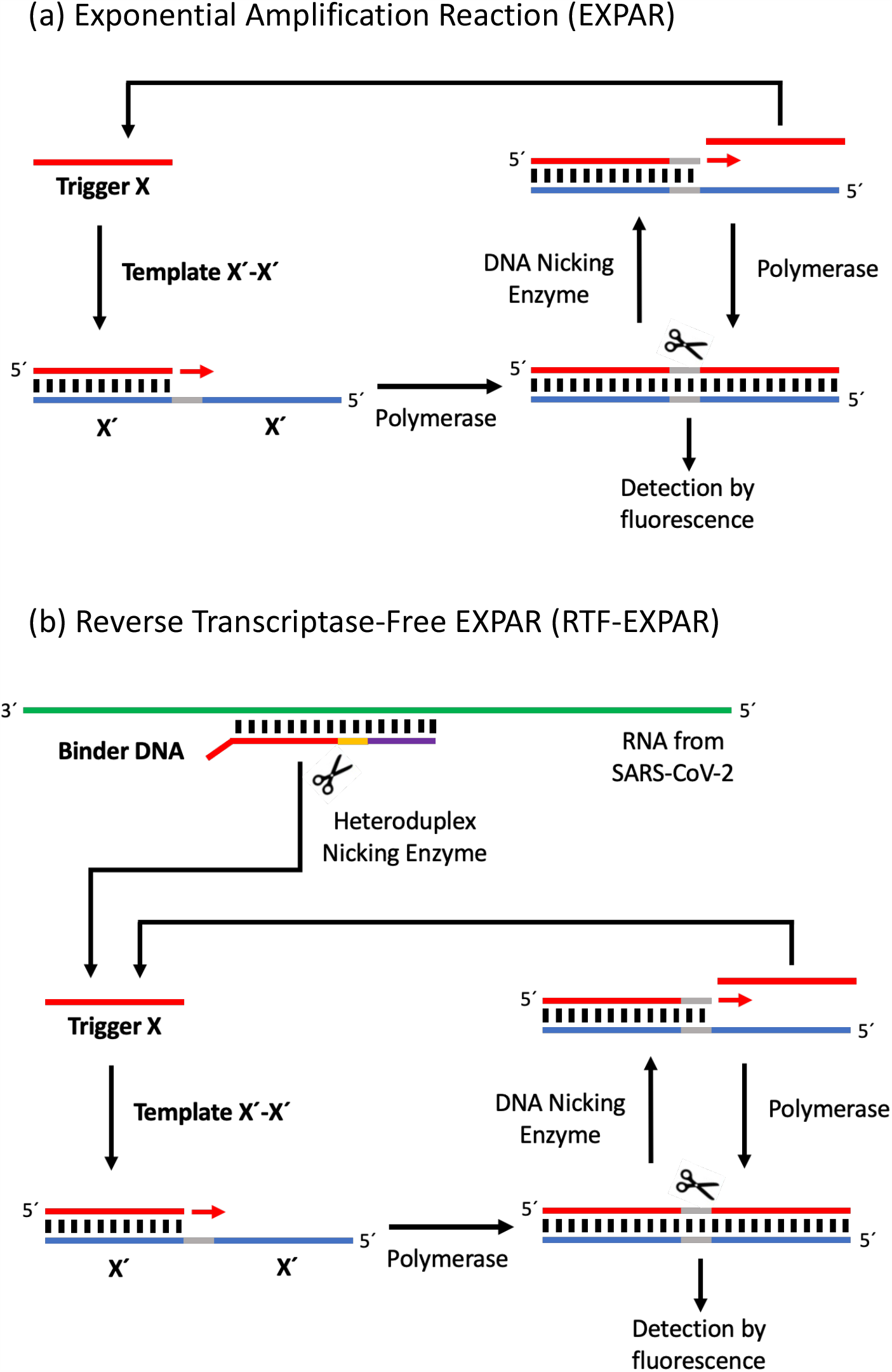
(a) Reaction Scheme for EXPAR: **Trigger X** anneals to **Template X’-X’** and is extended by a DNA polymerase (*Bst* 2.0 polymerase); the top strand of the newly formed duplex DNA is then cut by a nicking enzyme (Nt.*Bst*NBI); the released DNA (which is displaced by DNA polymerase in a subsequent extension reaction) is identical to **Trigger X** and is therefore able to prime another **Template X’-X’**. (b) Reaction Scheme for Reverse Transcriptase-Free EXPAR (RTF-EXPAR): **Binder DNA** anneals to viral RNA; the DNA strand of the DNA:RNA heteroduplex is cut by the restriction endonuclease *Bst*NI, which acts as a nicking enzyme by cutting the DNA strand only, the released DNA strand is **Trigger X**, which is then amplified by EXPAR.

## Data Availability

All relevant data is supplied in the text and supplementary information

## Acknowledgments

We would like to thank Dr A. Lovering for providing experimental equipment. We would also like to thank Public Health England for supplying the isolated patient RNA from SARS-CoV-2. This research was funded by the Midlands Integrative Bioscience Training Partnership funded by the Biotechnology and Bioscience Research Council (BB/R506175/1). There are no other competing interests.

## Author Contributions

T.R.D, M.R.H and J.H.R.T supervised the project; J.G.C, J-L.H.A.D, L.O.I, T.R.D, M.R.H and J.H.R.T designed the assay; A.D.B suggested the target gene and A.B. supplied the genomic material; I.R.C and C.D.S designed the analysis software; J.G.C undertook the research; J.G.C, T.R.D and J.H.R.T. wrote the manuscript.

## Experimental Section

### Materials

Milli-Q water purified with a Millipore Elix-Gradient A10 system (resistivity > 18 μΩ.cm, TOC ≤ 5ppb, Millipore, France) was used in all the experiments. Nt.*Bst*NBI, *Bst*NI and *Bst* 2.0 Polymerase were obtained from New England Biolabs (Hitchin, UK) as was the buffer, 10x Isothermal amplification buffer (200 mM Tris-HCl, 100 mM (NH_4_)_2_SO_4_, 500 mM KCl, 20 mM MgSO_4_, 1% Tween 20, pH 8.8) which was used in all the experiments. Superscript IV Reverse Transcriptase was obtained from ThermoFisher (Paisley, UK), DMSO (> = 99%) was obtained from Fisher Scientific (Loughborough, UK) and dsGreen 100x (an analogue of SYBR Green I), was obtained from Lumiprobe (Hannover, De). Bovine Serum Albumin (BSA, diluted to 4 mg/mL in water) and Single-Stranded Binding Protein (SSB, solution of 0.5 mgs in 20 mM Tris-HCl, pH 8.0, 0.5 M NaCl, 0.1 mM EDTA, 0.1 mM DTT, 50% Glycerol) was obtained from Sigma-Aldrich (Dorset, UK). All the nucleotide triphosphates and oligonucleotide sequences (desalted) were obtained from Sigma-Aldrich (Dorset, UK).

### PHE samples

All clinical specimens were handled in a Containment Level 2 laboratory. To prepare each sample, Viral Transfer Medium (VTM, 300 µL, Medical Wire ViroCult) from a nose and throat swab was added to Buffer AL (Qiagen) in a 1:1 ratio and heated to 60 °C for 30 minutes in a calibrated heat block. Samples were then extracted on the MagNAPure96 (Roche) automated extraction system and then run on the Abbott M2000 RT-qPCR Test for SARS-CoV-2 RNA Detection. For EXPAR assay development, positive and negative samples from the SARS-CoV-2 RNA assays were separately combined in MagNA Pure elution buffer (giving 29,080 RNA copies/µL for the combined positive sample). Upon receipt from PHE, each sample was diluted 400-fold with water, aliquoted into 50 µL vials and stored at −80 °C. Prior to use, each sample was submerged in ice and allowed to slowly melt; once melted the sample was used immediately before being cooled again for storage at −80 °C.

### Data Analysis and classification

To analyse the EXPAR real-time fluorescence amplification curves and data, a program in C# was developed. The program analyses the first 10 data points and calculates the mean value and standard deviation as a base line. Following generation of these two values, each subsequent data point is analysed to determine if its value minus the average value is greater than 10 standard deviations away from the mean. The cycle which meets this criterion is converted into a time and used as the minimum amplification time. Under the concentrations and conditions used in the RTF-EXPAR assay protocol, should the amplification time be less than 10 minutes, the output indicates the presence of SARS-CoV-2 RNA (true positive). For amplification times greater than 10 minutes, the output indicates a complete test and the absence of SARS-CoV-2 RNA (false positive).

### RTF-EXPAR Assay Protocol

The protocol first involves the preparation of three solutions, Part A, Part B and Part C, followed by an addition step and then finally an amplification step.

*Part A*:

1.50 µL of water, 2.50 µL of 10x Isothermal amplification buffer, 3.75 µL of BSA solution, 1.50 µL of *Bst* 2.0 DNA polymerase (1.6 U/µL), 0.75 µL of Nt.*Bst*NBI (10 U/µL).

*Part B*:

6.30 µL of water, 5.00 µL of 10x Isothermal amplification buffer, 0.75 µL of **Template X’- X’** (1 µM), 2.40 µL of MgSO^4^ (100 mM), 1.50 µL dNTP (10 nM), 0.75 µL of dsGreen (1:5 dilution in DMSO from 100x to 20x), 0.30 µL of SSB solution.

*Part C*:

(1) Sensitivity test (no RNA target): 3 µL of one trigger at **Trigger X** (100 nM, 10 nM, 1 nM, 100 pM, 10 pM, 1 pM and a blank)

OR

(2) Specificity test (no RNA target): 3 µL of one trigger at 100 nM (**Trigger X** or **Trigger A** or **Trigger B** or **Trigger C**) OR

(3) Reverse Transcriptase-Free EXPAR assay (two-pot RTF-EXPAR): 10 µL of RNA:DNA heteroduplex digestion mixture, prepared as follows: 25 µL of water, 5 µL of 10x Isothermal amplification buffer, 5 µL *Bst*NI (10 U/µL), 10 µL of **Binder DNA** (1 µM) and 5 µL of viral sample. The mixture is then incubated at 50 °C for 5 minutes.

OR

(4) Reverse Transcriptase-Free EXPAR assay (one-pot RTF-EXPAR): reagents are mixed together in the following order: 1 µL *Bst*NI (10 U/µL), 2 µL of **Binder DNA** (1 µM) and 3 µL of viral sample.

*Addition step*: Part B (17 µL) is added to a PCR tube, and to this is added Part C, followed by Part A (10 µL). The tube is then sealed, with the contents then subjected to amplification.

*Amplification step:* Isothermal incubation and fluorescence signal measurements are performed using an Agilent Mx3005P Real-Time PCR system (Didcot, UK) set to a constant temperature of 50 °C. The fluorescence is measured every 10 seconds over an incubation time of 30 minutes.

