## Supplementary Information for "Sub-5-minute Detection of SARS-CoV-2 RNA using a Reverse Transcriptase-Free Exponential Amplification Reaction, RTF-EXPAR"

Contents:

1. EXPAR Oligonucleotide Sequences Page S2
2. EXPAR Sensitivity and Specificity Tests Page S3
3. RTF-EXPAR Assay Data Page S5

### 1. EXPAR Oligonucleotide Sequences

Supplementary Table 1: Oligonucleotides used during EXPAR.

| Name | Sequence (5' – 3') |
|---|---|
| **Trigger X** | AGG GTA AAC CAA ATA CC |
| **Trigger A** | AGG GTT AAA CCA CCG CC |
| **Trigger B** | AGG GTC CTT AAC TTG CC |
| **Trigger C** | CCG GGA TTG GTT GAT |
| **Template X′-X′** | GGT ATT TGG TTT ACC CTG TGA GAC TCT GGT ATT TGG TTT ACC CT |
| **Binder DNA** | **AGG GTA AAC CAA ATA CC**T GGT GTA TAC GTT |

Key:
Magenta – Non-binding fragment
Blue – *Bst*NI recognition site (complementary to required 5'-CCAGG-3'sequence in RNA target)
**Bold** – **Trigger X** sequence in **Binder DNA** sequence

### 2. EXPAR Sensitivity and Specificity Tests

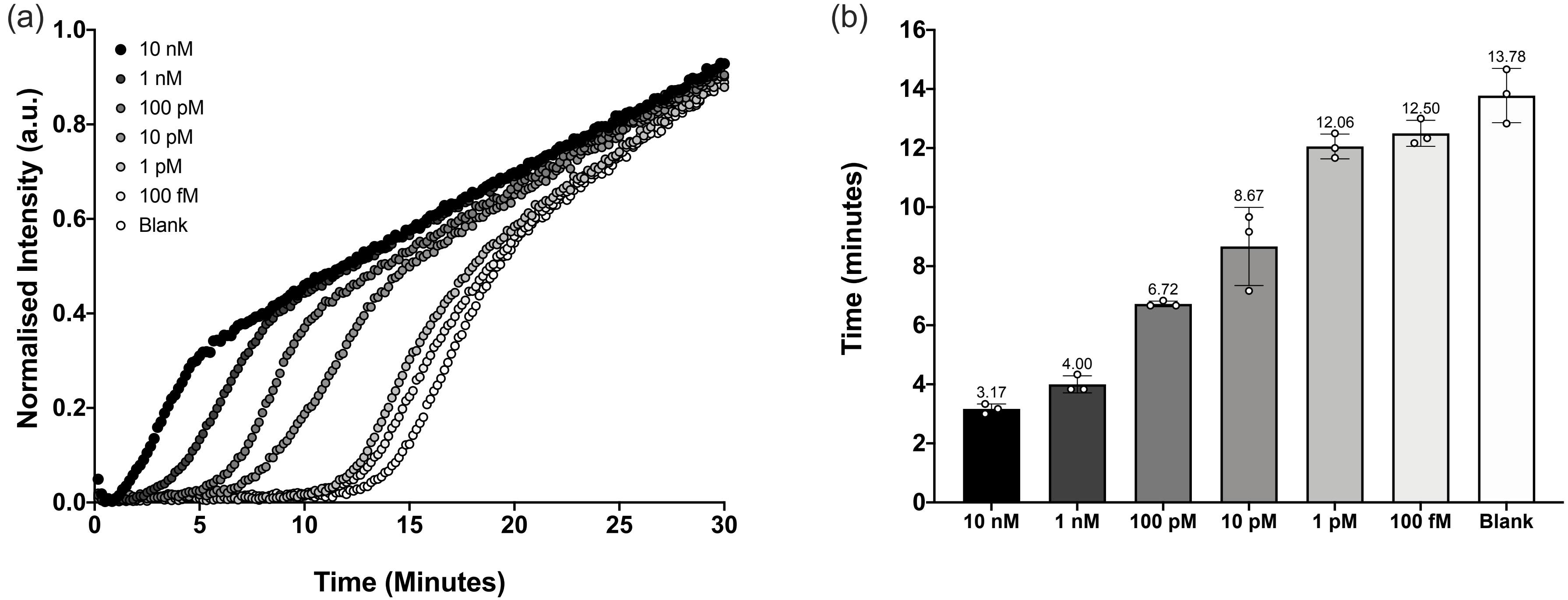


**Supplementary Figure 1**. EXPAR sensitivity test data showing: (a) normalised average fluorescence intensity data (a.u) plotted against time (min) and (b) mean amplification times to greater than 10 standard deviations from the baseline fluorescence, plotted against **Trigger X** concentration (10 nM, 1 nM, 100 pM, 10 pM, 1 pM, 100 fM and a blank) in the presence of **Template X'-X'** (25 nM). Runs performed in triplicate (n = 3). Error bars in datasets are the standard deviations of the 10-sigma time. For other reagents and conditions, see RTF-EXPAR protocol.

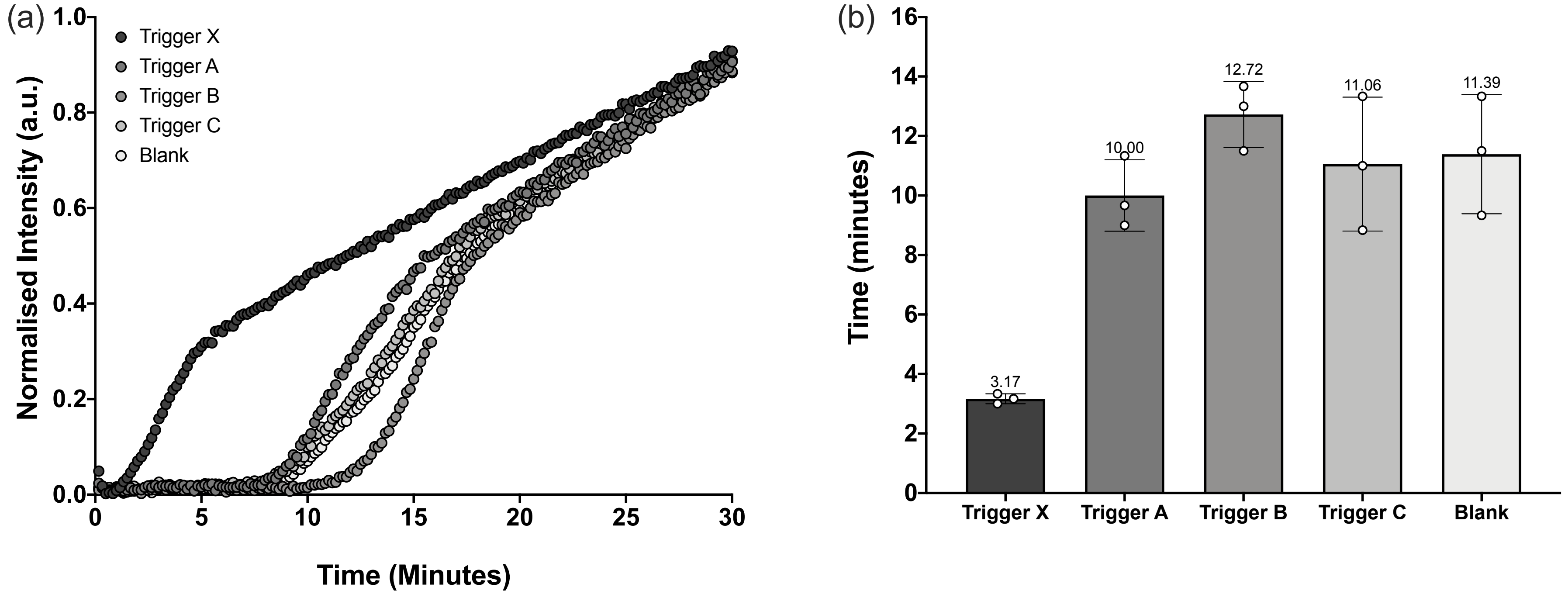


**Supplementary Figure 2**. EXPAR specificity test data showing: (a) normalised average fluorescence intensity data (a.u) plotted against time (min) and (b) mean amplification times to greater than 10 standard deviations from the baseline fluorescence signal, plotted against **Trigger X** (fully complementary), **Trigger A** (non-complementary), **Trigger B** (non-complementary), **Trigger C** (non-complementary) and a blank (no trigger). **Template X´-X´** concentration 25 nM, trigger concentration 10 nM. Runs performed in triplicate (n = 3). Error bars in datasets are the standard deviations of the 10-sigma time. For other reagents and conditions, see RTF-EXPAR protocol.

### 3. RTF-EXPAR Assay Data

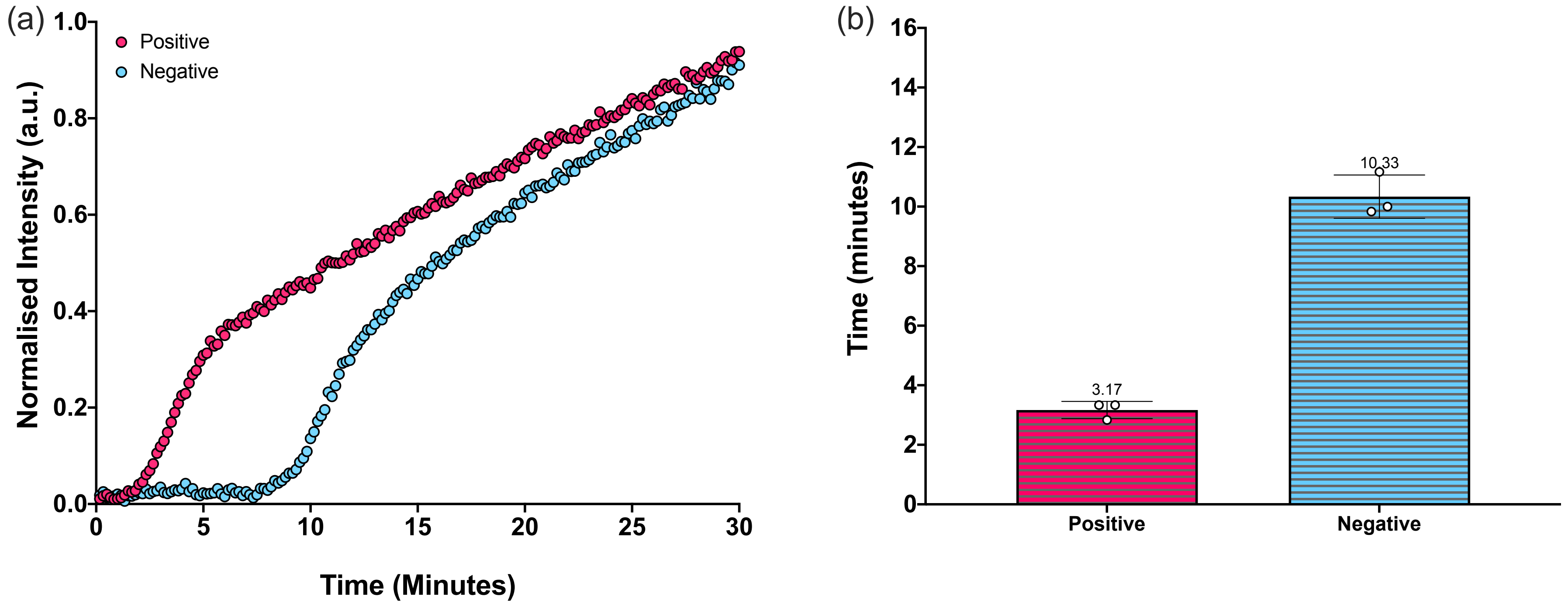


**Supplementary Figure 3**. Two-Pot RTF-EXPAR data showing: (a) normalised average fluorescence intensity data (a.u) plotted against time (min) and (b) mean amplification times to greater than 10 standard deviations from the baseline fluorescence signal, plotted against positive (72.7 copies/μL) and negative COVID-19 RNA patient samples. Runs performed in triplicate (n = 3). Error bars in datasets are the standard deviations of the 10-sigma time. For other reagents and conditions, see RTF-EXPAR protocol.

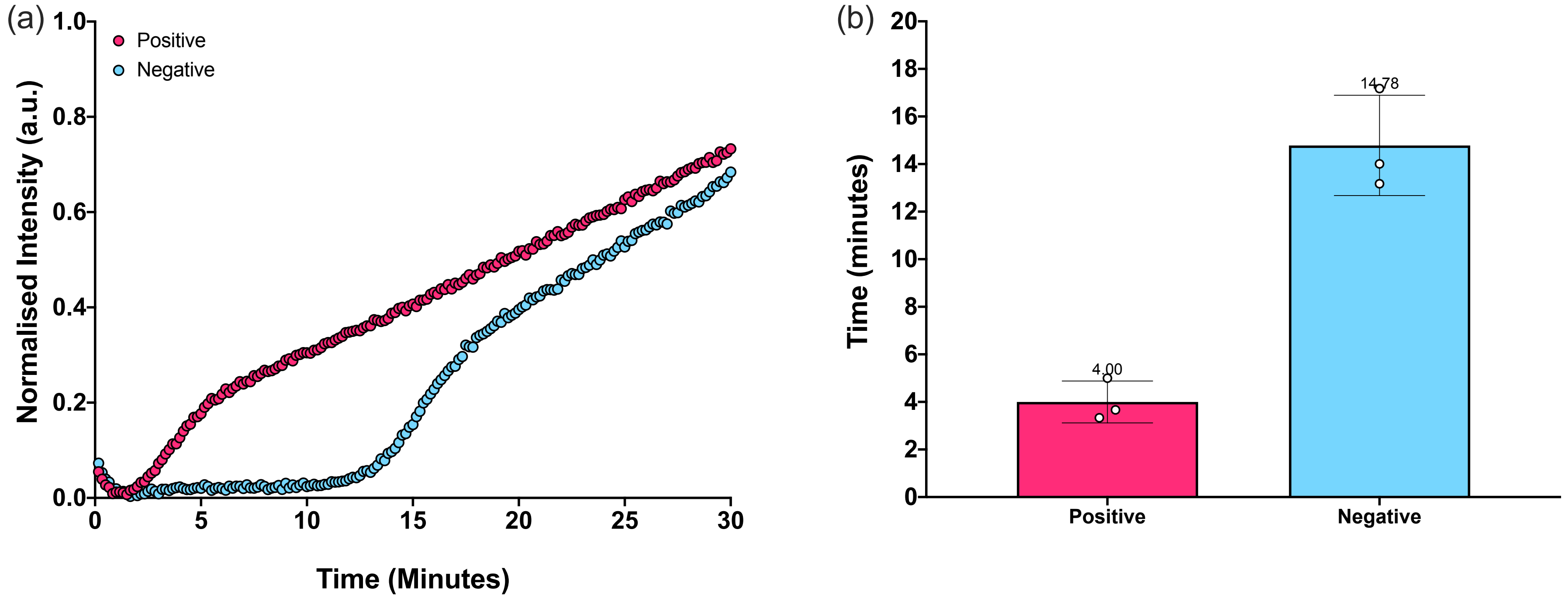


**Supplementary Figure 4**. One-Pot RTF-EXPAR data showing: (a) normalised average fluorescence intensity data (a.u) plotted against time (min) and (b) mean amplification times to greater than 10 standard deviations from the baseline fluorescence signal, plotted against positive (72.7 copies/µL) and negative COVID-19 RNA patient samples. Runs performed in triplicate (n = 3). Error bars in datasets are the standard deviations of the 10-sigma time. For other reagents and conditions, see RTF-EXPAR protocol.

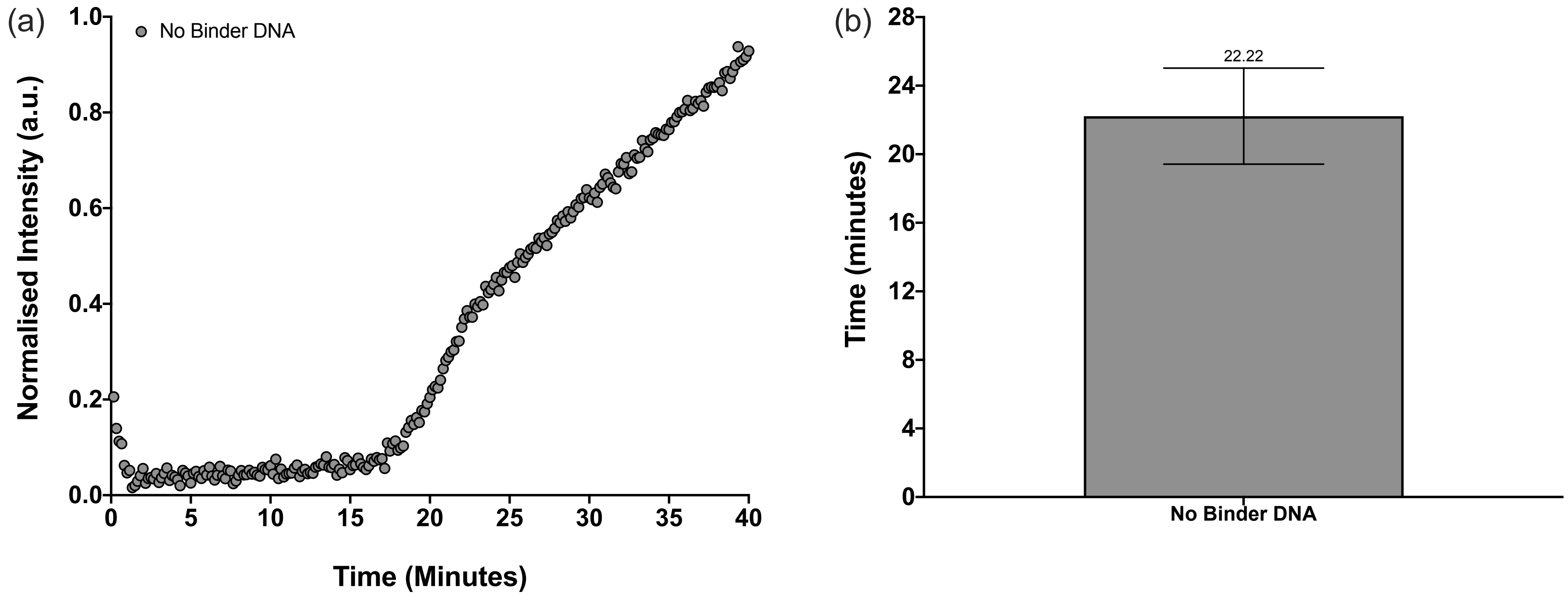


**Supplementary Figure 5**. One-Pot RTF-EXPAR data showing: (a) normalised average fluorescence intensity data (a.u) plotted against time (min) and (b) mean amplification time to greater than 10 standard deviations from the baseline fluorescence signal, plotted against positive (72.7 copies/μL) COVID-19 patient samples in the absence of **Binder DNA**. Runs performed in triplicate (n = 3). Error bars in datasets are the standard deviations of the 10-sigma time. For other reagents and conditions, see RTF-EXPAR protocol.

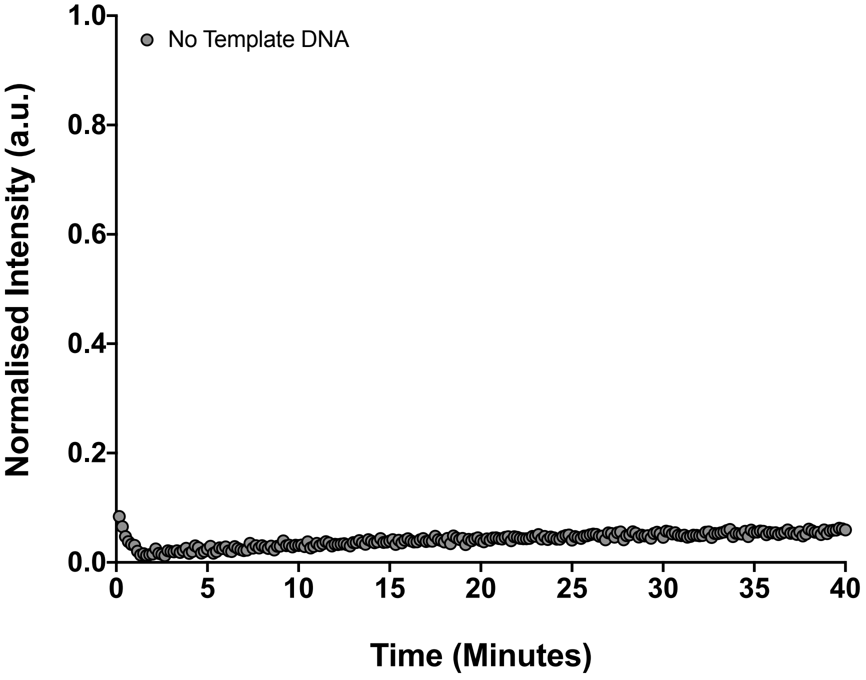


**Supplementary Figure 6**. One-Pot RTF-EXPAR data showing normalised average fluorescence intensity data (a.u) plotted against time (min) for positive (72.7 copies/µL) COVID-19 patient samples in the absence of **Template X´-X´**. Runs performed in triplicate (n = 3). For other reagents and conditions, see RTF-EXPAR protocol.

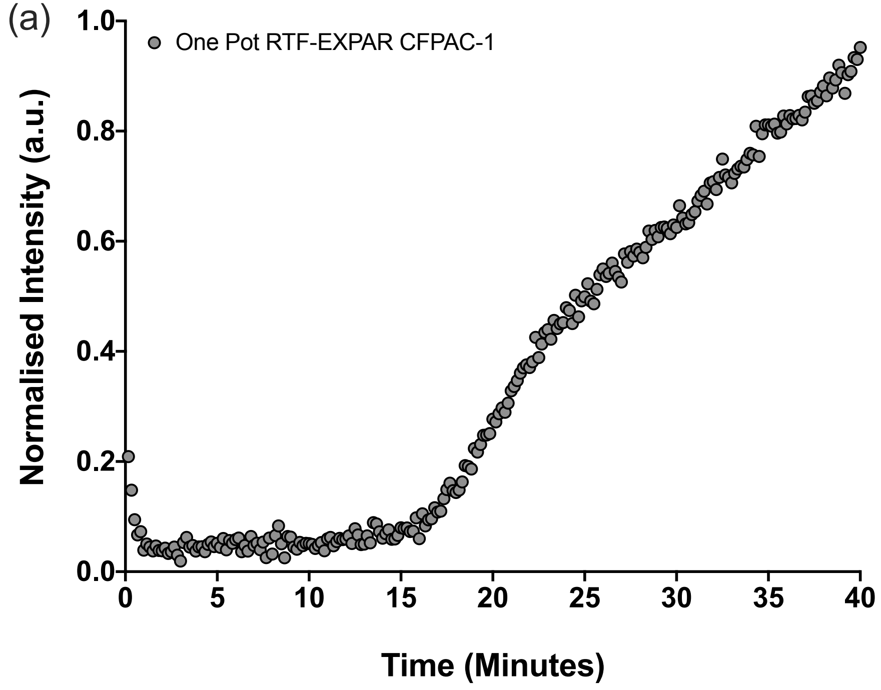


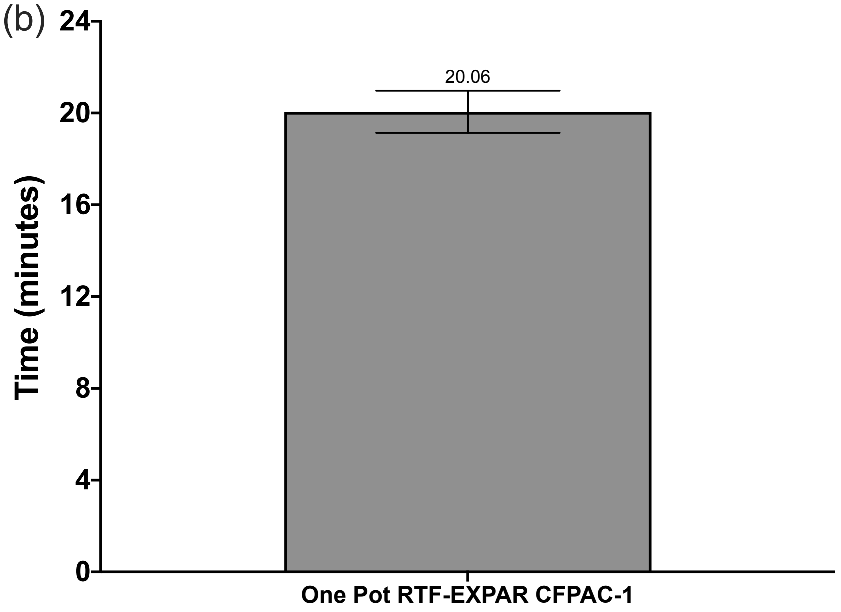


**Supplementary Figure 7**. One-Pot RTF-EXPAR data showing: (a) normalised average fluorescence intensity data (a.u) plotted against time (min) and (b) mean amplification time to greater than 10 standard deviations from the baseline fluorescence signal, plotted against sample containing RNA isolated from the CFPAC-1 human ductal pancreatic adenocarcinoma cell line (160.3 ng/μL). Runs performed in triplicate (n = 3). Error bars in datasets are the standard deviations of the 10-sigma time. For other reagents and conditions, see RTF-EXPAR protocol.
